# Quarantine and testing strategies in contact tracing for SARS-CoV-2: a modelling study

**DOI:** 10.1101/2020.08.21.20177808

**Authors:** Billy J Quilty, Samuel Clifford, Stefan Flasche, Adam J Kucharski, CMMID COVID-19 Working Group, W John Edmunds

## Abstract

**Background:** In most countries, contacts of confirmed COVID-19 cases are asked to quarantine for 14 days following exposure, to limit asymptomatic onward transmission. We assessed the merit of RT-PCR testing in reducing the length of quarantine, using the UK as a case study.

**Methods:** We used an agent-based model to simulate an exposed contact’s contact tracing detecting delay, incubation period, probability to become symptomatic, infectivity profile, and time-varying PCR detection probability. Assuming self-isolation on symptom onset, we assess the impact of the current 14 day quarantine strategy for all exposed contacts on their onward transmission potential and compare to alternative quarantine lengths with or without PCR tests during quarantine.

**Findings:** Self-isolation on symptoms onset alone can prevent 39% (95% Uncertainty Interval for super spreading events: 34, 45%) of onward transmission from secondary cases. An additional 14 days post-exposure quarantine for all contacts reduces transmission by 70% (95% UI: 39, 90%). A negative PCR test taken upon tracing or 7 days after exposure can reduce transmission by 62% (95% UI: 40, 84%) or 68% (95% UI: 40, 88%) respectively. Halving contact tracing delays of currently 4 days reduces pre-tracing transmission potential from 26% (95% UI: 7, 56%) to 14% (95% UI: 5, 42%).

**Interpretation:** PCR testing may allow for a substantial reduction in quarantine needs or even replacing quarantine requirements with no or a small excess in transmission risk respectively. Reducing contact tracing delays can help prevent a substantial amount of transmission.

**Funding:** National Institute for Health Research, UK Research and Innovation, Wellcome Trust, and EU Horizon 2020.

**Research in context:** *Evidence before this study:* During the SARS-CoV-2 pandemic, a standard 14-day quarantine period has been required from the day a contact was exposed to an index case to avert onwards transmission. This approach aims to avoid infected contacts returning to their normal life during their pre-symptomatic period. This strategy presents a crucial part in the global pandemic response to interrupt transmission chains, however it places considerable social and economic pressure on contacts. As case numbers are rapidly rising across Europe an effective, practical and widely accepted strategy to limit secondary transmission is key to the control of SARS-CoV-2. To the best of our knowledge, this is the first analysis of optimal strategies to reduce transmission from contact traced secondary cases.

*Added value of this study:* We simulated the ability of different quarantine and testing strategies to reduce the transmission potential of secondary cases. We found that the common post-exposure quarantine period of 14 days is equally effective as PCR testing of contacts after 7 days of quarantine. PCR testing immediately on tracing with no quarantine requirements following a negative test result would avert slightly less transmission potential from secondary cases (62% vs 70%). Delays in identifying contacts and hence delayed quarantine as well as low adherence substantially reduces the ability to reduce secondary transmission.

*Implications of all the available evidence:* The ability to test timely and at scale could shorten current delays in contact tracing and thereby reduce the risk for secondary transmission. Similarly, such testing capacity would allow to substantially shorten necessary quarantine periods and dampen the economic and social impact while potentially increasing compliance.

## Introduction

In order to break transmission chains of SARS-CoV-2, testing of cases and the tracing and quarantine of their contacts has been employed as key non-pharmaceutical intervention (NPI) in many countries. This measure aims to prevent onward transmission from secondary infections (individuals infected by an index case). This method has been employed successfully to prevent new outbreaks in countries such as South Korea without the need for “lockdown”-style measures. Current guidance in England is that traced individuals must self-isolate from the moment they are traced until 14 days have elapsed from their exposure to the index case. This 14 days represents the upper bound for the incubation period (1), when >95% of eventually-symptomatic persons will have developed symptoms and subsequently enter a further period of self-isolation (10 days in England). At ∼5 days, 50% of eventually-symptomatic individuals will have developed symptoms, as well as reach the peak probability of detection by PCR testing (2). As such, it is possible that testing traced contacts may detect latent and asymptomatic cases, allowing for a reduction in the 14-day post-exposure period. Key to this is the timing of testing, as testing contacts too early may result in false-negative results. Another crucial factor is the delays in testing and tracing, i.e, how long has passed since exposure of the index case to the isolation of their contacts, as approximately half of SARS-CoV-2 transmission occurs before the onset of symptoms (3). Additionally, there is evidence that the current 14 day period is poorly adhered to, with only 10.9% of contacts of cases reportedly not leaving the house in the 14 days following exposure to the index case (4). It is possible that reducing this quarantine period may increase adherence and therefore avert more transmission overall.

We used the incubation period (1) and infectivity profile (3,5) of SARS-CoV-2 combined with the observed delays in testing and tracing observed in England in late-August 2020 to evaluate a range of quarantine and testing strategies for contacts of cases identified through contact-tracing, varying: the required post-exposure quarantine period, the timing of tests, and the number of tests. We estimate the amount of transmission potential averted in each strategy which would otherwise be spent in the community. We also investigate the effect of reducing testing and tracing delays, as well as the impact of reduced adherence to quarantine.

## Method

### Contact tracing model

We used a stochastic, individual-based model to determine the effect of different quarantine and testing strategies on the transmission potential of secondary cases, accounting for variation in exposure timings, incubation period, and test sensitivity. Following the notation of Kretzschmar et al. (2020) (6), we consider the following events to be relevant to the tracing of the contacts of an index case - an individual assumed to be newly-symptomatic with COVID-19 and seeking a test (Figure 1). Each of the following variables are specific to an individual, but we omit a subscript, *i*, for brevity. An individual is exposed and becomes infected at time *T* _0_. We assume the index case has onset of symptoms at time *T* _2_, lasting until time *T* _2_′. For sensitivity, we assume testing of the index case occurs at time *T* _3_, 1, 2 or 3 days after symptom onset, with the results of the test available at time *T* _3_′ ; those testing positive will go on to isolate for 10 days from their symptom onset (7). We assume that positive tests are immediately referred to contact tracers, with the index case’s contacts’ information sourced at *T* _4_, and these contacts are then traced and quarantined at time *T* _4_′. For comparison, we consider the baseline for index case testing delay to be 2 days.

**Figure 1.**
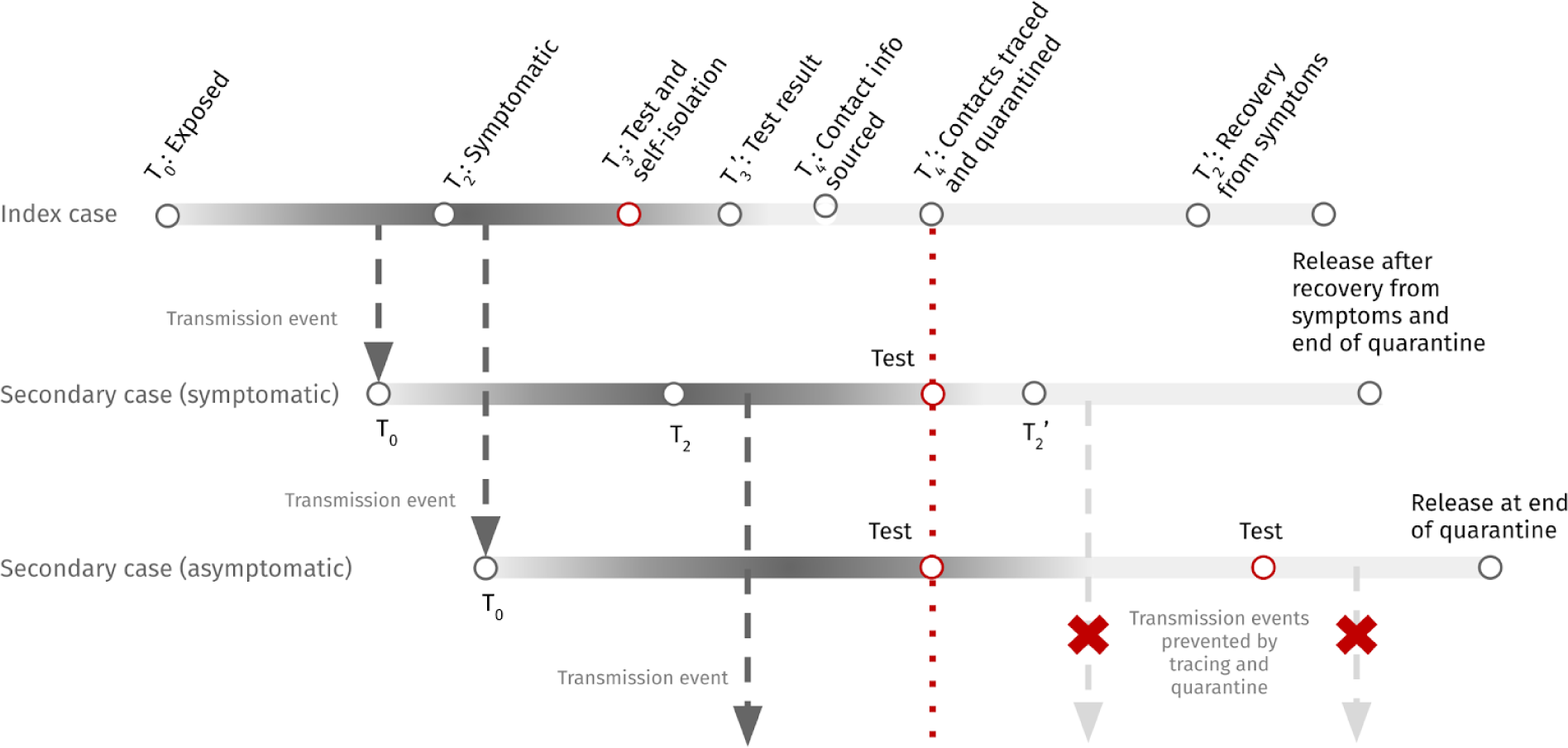
Example schematic of the contact tracing process and associated timings where an index case causes two secondary cases, one of which is symptomatic and one of which is asymptomatic. Darker shaded regions of each cases’ timeline indicate periods of increased infectivity. Arrows indicate transmission events, with red crosses indicating transmission prevented through quarantine of traced contacts. The symptomatic and asymptomatic secondary cases are traced and quarantined (dashed red line), and in the two-test scenario shown, receive a test at this time as well as at a fixed number of days since their exposure (red circles). Secondary cases are then released from quarantine after the quarantine time has elapsed and if they no longer show symptoms.

Rather than assuming a specific time at which the infectious period begins (*T* _1_) we consider the infectivity profile, i.e, a distribution of times at which transmission is likely to occur. This distribution is derived by considering the incubation period (time from exposure to onset of symptoms), *T* _2_ − *T* _0_, and delay from infectiousness to onset of symptoms *T* _2_ − *T* _1_, and using the (corrected) infectivity profile from He et al. (2020) (5) with incubation periods sampled from the distribution in (1).

We parameterise the delays associated with the contact tracing system (having a test to receiving the results (*T* _3_′ − *T* _3_), sourcing contact information (*T* _4_ − *T* _3_′), and tracing (*T* _4_′ − *T* _4_)) according to available NHS Test and Trace data from the week 20 August 2020 to 26 August 2020 (Table 2) (8). These times are reported as 24 hour intervals ({0 ≤ *t* < 1, 1 ≤ *t* < 2, 2 ≤ *t* < 3, *t* ≥ 3} days), which we used to derive a Gamma distribution considering the delay in each index case’s awaiting a test result, sourcing of contacts and tracing of contacts as doubly-censored observations on the specified time intervals using the fitdistcens function from the fitdistrplus package in R (9). For sensitivity analysis, we halve the sampled delays from the Test and Trace system to determine the impact of improving throughput.

**Table 1.**
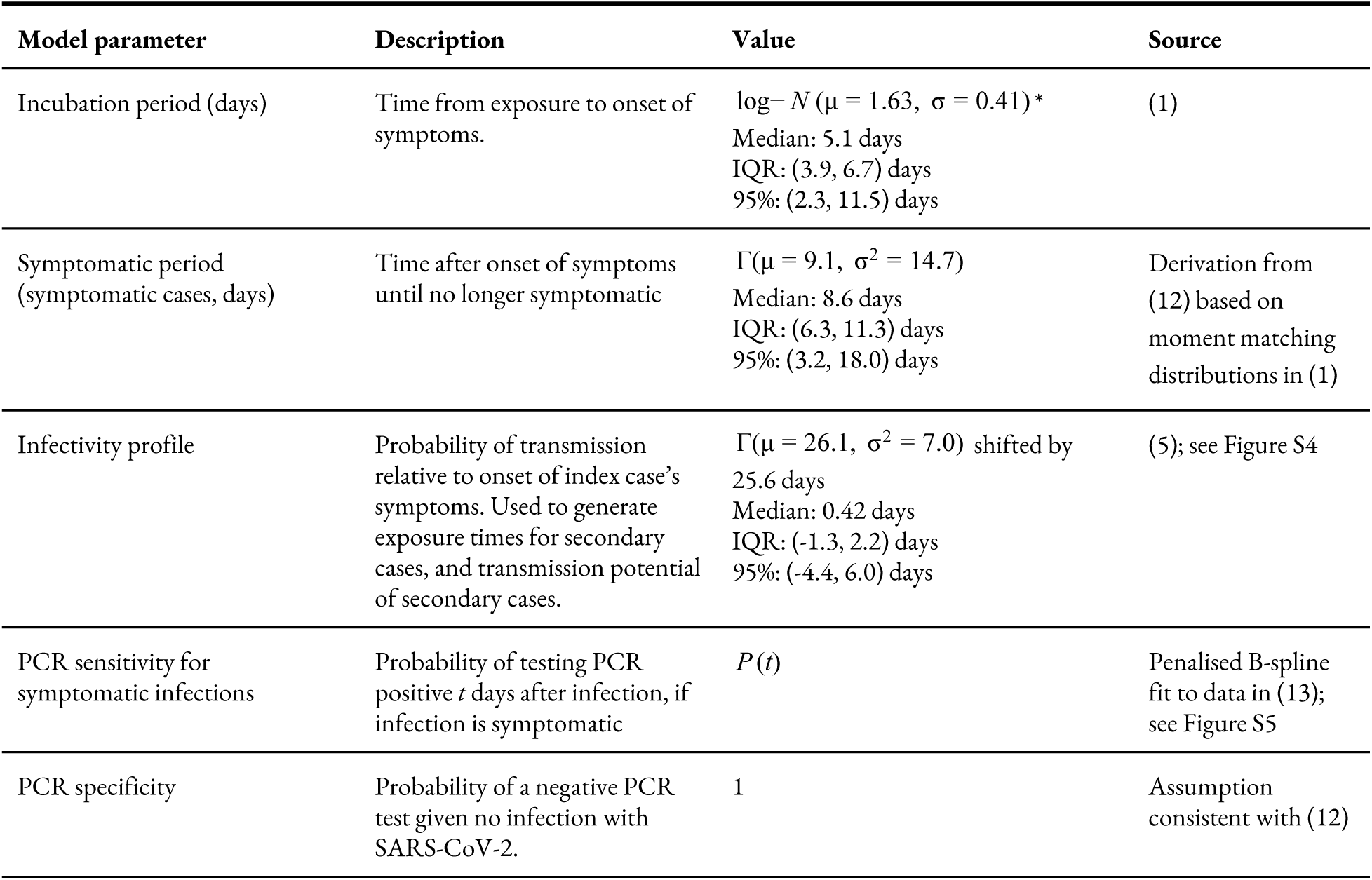
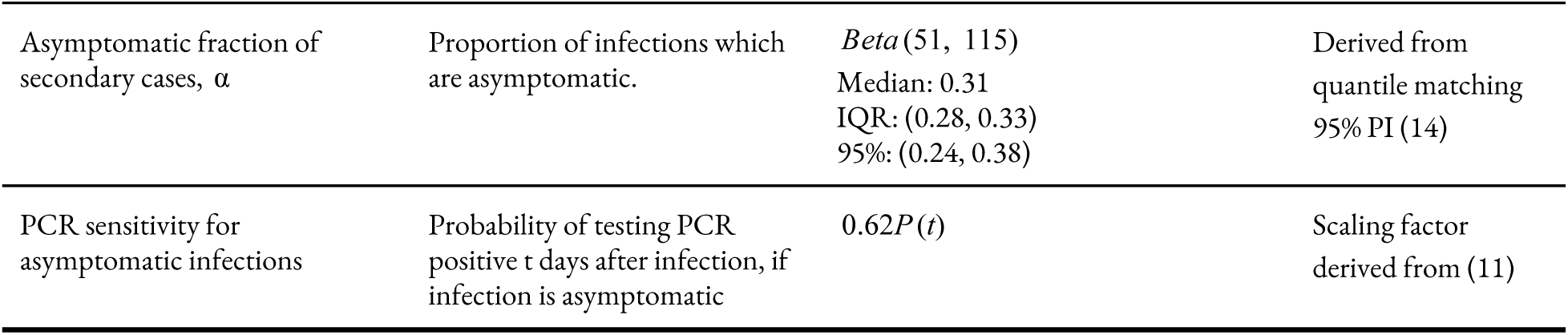
Values of parameters in simulation of cases’ infection histories and PCR testing. Gamma distributions are parameterised in terms of a mean and variance, Γ(μ, σ^2^), and these are converted to shape and rate parameters via moment matching. Where quantiles are given but no distribution described, the parameter is derived from other distributions in the table and has no closed-form. *Parameters are location and scale for log-Normal distribution, not summary statistics of observed incubation period.

**Table 2:** Time periods for return of index cases’ test result, sourcing of contacts, and tracing of contacts derived from NHS Test and Trace for the week 20 August 2020 - 26 August 2020 (8). Gamma distribution mean, standard deviation, median and 95% prediction interval for time to source contacts from case and time to trace said contacts. *The total is derived from the sum of 100,000 sampled values and no closed form distribution exists.

| Delay distribution | Mean, $\mu$ | Std. Dev., $\sigma$ | 2.5% | 50% | 97.5% |
| --- | --- | --- | --- | --- | --- |
| Return of index cases' test result | 2.58 days | 2.22 days | 0.14 days | 1.97 days | 8.34 days |
| Sourcing of contacts | 0.74 days | 0.91 days | 0.00 days | 0.42 days | 3.29 days |
| Tracing of contacts | 0.61 days | 0.66 days | 0.01 days | 0.39 days | 2.41 days |
| Total* | 3.93 days | 2.49 days | 2.13 days | 3.39 days | 10.18 days |

### Quarantine and testing strategies

We consider periods between 0 and 14 days after exposure to the index case for which contacts must quarantine until, reflecting current UK guidance (10). The currently recommended duration, 14 days since last exposure to the index case, represents the time by which it is expected that >95% of ever-symptomatic cases will display symptoms (1) and continue to self-isolate; however it should be noted that cases only enter quarantine when they are traced, and hence there is a pre-quarantine period where secondary cases may be infectious.

We also consider zero, one, or two PCR test strategies. The zero test scenarios represent a quarantine-only strategy. The one test strategy sees individuals tested at the end of their specified quarantine period (defined as when *t* days have passed since exposure) and released from quarantine two days later if negative (based on reported UK data, Table 2).. If cases are traced after the date of their specified test, we assume a test occurs upon tracing. The two test strategy sees individuals additionally tested at the time that they are initially traced and enter quarantine. If the difference between the test administered on tracing and the test at the end of quarantine is less than the assumed delay in getting a test result, we assume that the initial test upon tracing is not conducted. Any secondary case displaying symptoms at any point post-exposure will continue to isolate until 10 days have passed since onset of symptoms (7). We assume that asymptomatic secondary cases never develop symptoms and hence will not self-isolate. We assume that asymptomatic persons have 62% the probability of detection at any given point in their infection compared to a symptomatic case (11). We assume the same basic transmission potential for asymptomatic and symptomatic individuals. Further details on the testing scenarios, infection history generation, and test sensitivity are provided in Table 1.

### Outcomes measured

For each secondary case we calculate the mass of the infectivity profile distribution from exposure to post-tracing isolation as a measure of transmission potential prior to quarantine. Similarly, the mass of the infectivity after release is a measure of transmission potential after quarantine, parameterised in terms of number of days from onset (3). We assume that cases cannot transmit before their exposure, and hence limit the transmission potential to be positive and rescale to account for this.

To investigate the effect of reduced or waning adherence, we weight the infectivity profile by a function, *w*(*t*), which represents the probability that each secondary case is adhering to quarantine guidance. Our baseline case, perfect adherence, is *w*(*t*) ≡ 1. We consider reduced adherence for sensitivity, from *w*(*t*) ≡ 0.9 to *w*(*t*) ≡ 0.1, with 0.1 representing the observed rate of adherence for contacts of cases in the UK (average 10.9%) (4). The amount of transmission potential during quarantine or isolation, between times *t*_1_ and *t*_2_, then, is non-zero and represents the effect of imperfect compliance and adherence at a time since the initial quarantine, *t*_0_ :

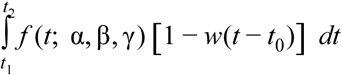

where *f* is the Gamma probability density function representing the infectivity profile with shape, α, rate, β, and shift, γ, Where an individual develops symptoms after their release from quarantine, we assume that they return to self-isolation until they are no longer symptomatic and at least 10 days have passed since their onset of symptoms, as well as the cessation of symptoms. Adherence continues according to the same function of time since initial quarantine, *t*_0_, rather than restarting when the individual re-enters isolation. This scaling of the infectivity profile, rather than sampling which secondary cases comply and the point at which complying cases stop adhering, reflects an interest in expected transmission potential reduction across simulations rather than an attempt to characterise how many additional infections each secondary case goes on to cause.

The amount averted due to quarantine and testing is then 1 minus the sum of the transmission potential occurring prior to quarantine, during quarantine (if adherence is not perfect), and after quarantine (including that period spent in self-isolation if symptomatic). Assuming that the majority of SARS-CoV-2 transmission is driven by superspreading events (15), we report the uncertainty associated with the average secondary transmission potential averted per super spreading event by simulating 1000 index cases with 10 secondary cases and calculate the median and 50%, 95% quantiles for the secondary transmission potential averted per index case. We also report the median and 95% uncertainty interval of these simulated values based on 1000 simulated index cases each with 100 secondary cases generated in the supplement. These represent the variability of individual level reductions and span the full range from 0% to 100% reduction in transmission potential in many cases (Figure S6).

The model was coded in R and is available at https://github.com/cmmid/pcr_test_trace

## Results

### Tracing delays

The summary statistics of the fitted distributions of return of index cases’ test results, and subsequent sourcing and tracing of contacts, are given in Table 2 and Figure S3. A majority of all activities relevant to contact tracing are completed within 24 hours of their beginning. The average modelled contact tracing takes approximately 3.9 days from time of initial test to completion of tracing. Here we have assumed that the duration of each of these activities are independent.

### Reduction in transmission potential

The effect of longer quarantine periods is to avert a greater proportion of transmission potential spent in the community (Figure 2). Relying only on symptomatic persons strictly self-isolating upon developing symptoms, the transmission potential averted after a 0-day quarantine is 39% (50% UI: 39%, 39%; 95% UI: 34%, 45%). The amount of transmission potential averted rises to 56% (50% UI: 43%, 66%; 95% UI: 34%, 77%) at 7 days post-exposure, and 70% (50% UI: 56, 80%; 95% UI: 39%, 90%) at 14 days. The uncertainty in these estimates is due to secondary cases’ time of exposure being widely distributed around an index case’s onset, as well as the assumption that symptomatics begin a new 10-day period of self-isolation upon symptom onset (which does not occur for asymptomatics) (Figure 2). As such, the majority of the risk comes from asymptomatic persons, most notably for shorter quarantine periods.

**Figure 2:**
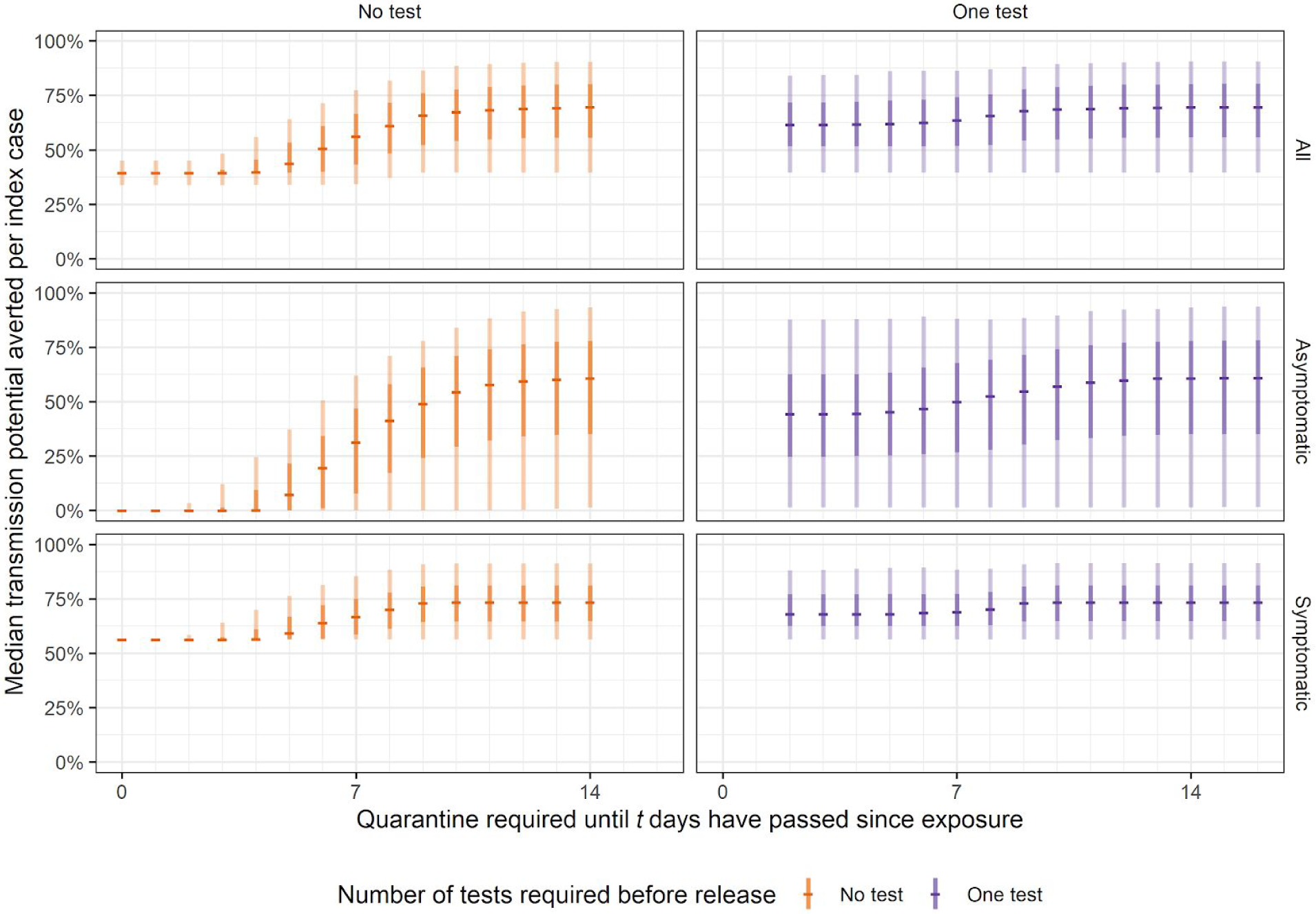
Transmission potential averted (integral of infectivity curve over time spent in quarantine and post-quarantine isolation) in the no test and one test scenarios, stratified by type of infection (all, asymptomatic, and symptomatic). Central bars indicate the median amount of transmission potential averted for a required time since exposure for which traced individuals must quarantine until. 95% and 50% uncertainty intervals indicated by light and dark shaded bars, respectively.

### Effect of PCR testing

The amount of transmission potential averted can be increased if PCR testing is conducted on the final day of quarantine (or upon tracing, if the specified quarantine period ends before a case is traced). The introduction of an immediate test with a two day turnaround (effectively a two-day quarantine) is to avert 62% (50% UI: 52%, 72%; 95% UI: 40%, 84%) of infectivity (Figure 2). However as the quarantine period increases, the relative contribution of a test is lessened. In the maximum stringency scenarios, with 14 days of mandatory quarantine, approximately 70% of transmission potential is averted both with and without a test (Figure 2, Table 3). The additional benefit of another test upon tracing (in addition to the end-of-quarantine test) is minor for longer quarantine periods (Figure S1, Table 3), however, it may have utility for rapid tracing of the contacts of secondary cases.

**Table 3:**
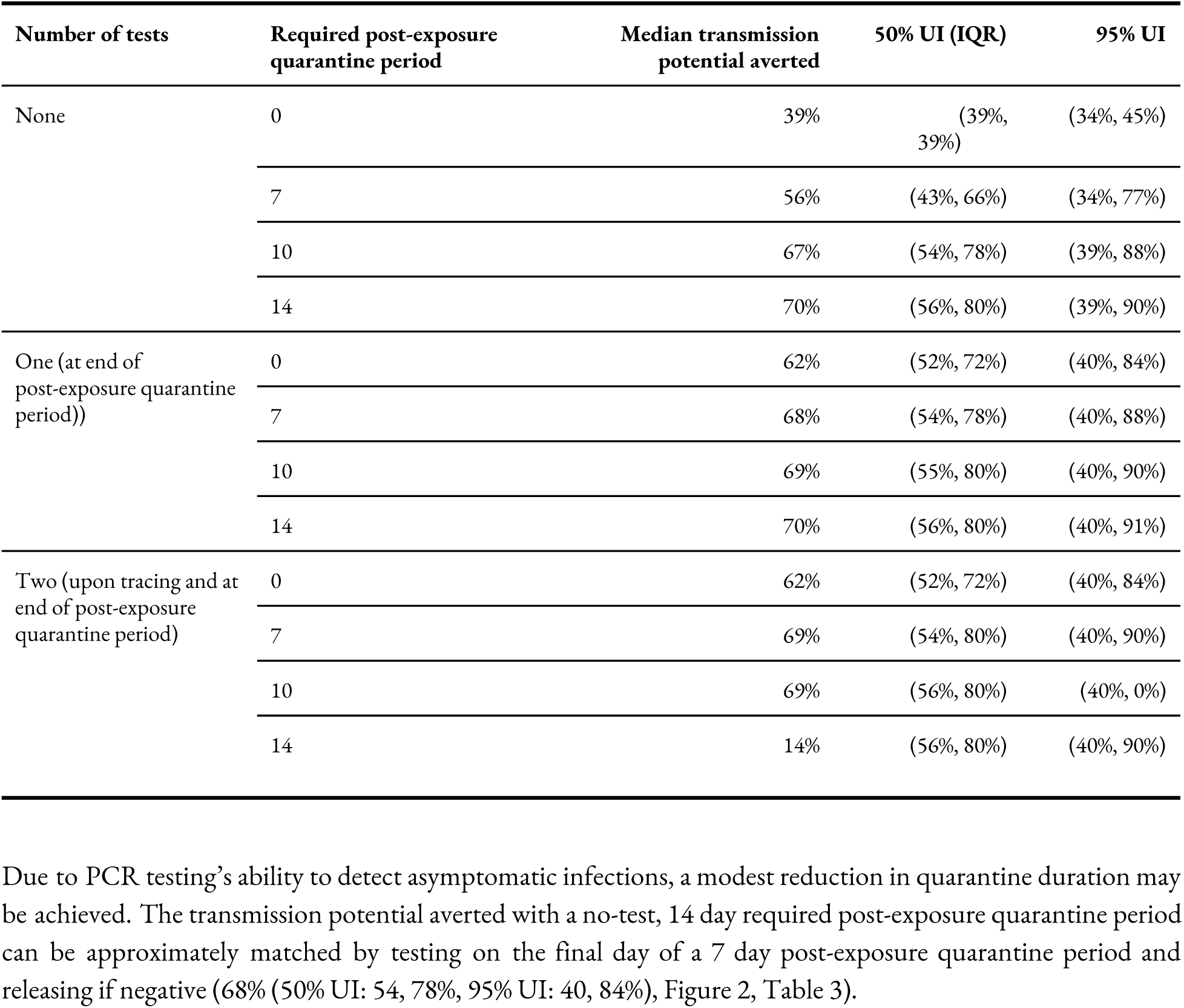
Transmission potential of secondary cases averted, stratified by the required number of days post-exposure to the index case that secondary cases must isolate until, and the number of tests conducted, with observed delays (Table 2). Note: with post-exposure quarantine periods shorter than the observed delays, we assume that a test takes place as soon as the case is traced. Similarly, we assume cases quarantine from the time of tracing up until the end of this period, so a 14-day post-exposure period does not represent 14 days in quarantine due to delays in testing and tracing.

| Number of tests | Required post-exposure quarantine period | Median transmission potential averted | 50% UI (IQR) | 95% UI |
| --- | --- | --- | --- | --- |
| None | 0 | 39% | (39%, 39%) | (34%, 45%) |
|  | 7 | 56% | (43%, 66%) | (34%, 77%) |
|  | 10 | 67% | (54%, 78%) | (39%, 88%) |
|  | 14 | 70% | (56%, 80%) | (39%, 90%) |
| One (at end of post-exposure quarantine period)) | 0 | 62% | (52%, 72%) | (40%, 84%) |
|  | 7 | 68% | (54%, 78%) | (40%, 88%) |
|  | 10 | 69% | (55%, 80%) | (40%, 90%) |
|  | 14 | 70% | (56%, 80%) | (40%, 91%) |
| Two (upon tracing and at end of post-exposure quarantine period) | 0 | 62% | (52%, 72%) | (40%, 84%) |
|  | 7 | 69% | (54%, 80%) | (40%, 90%) |
|  | 10 | 69% | (56%, 80%) | (40%, 0%) |
|  | 14 | 14% | (56%, 80%) | (40%, 90%) |

Tests act mostly to reduce the transmission potential of asymptomatic individuals, who would not otherwise be detected; for example, a test upon tracing with 0 days quarantine results in a median 44% (50% UI: 25, 62%; 95% UI: 1, 88%) of an asymptomatic persons’ transmission potential averted, as opposed to 0% without a test (Figure 2).

Due to PCR testing’s ability to detect asymptomatic infections, a modest reduction in quarantine duration may be achieved. The transmission potential averted with a no-test, 14 day required post-exposure quarantine period can be approximately matched by testing on the final day of a 7 day post-exposure quarantine period and releasing if negative (68% (50% UI: 54, 78%, 95% UI: 40, 84%), Figure 2, Table 3).

### Reducing tracing delays

Longer delays from the index case’s symptom onset to tracing of secondary cases increase the transmission potential of as yet untraced secondary cases in the community. As this occurs prior to tracing, it is independent of the specified duration of quarantine or number of tests conducted. At current observed delays from onset to tracing (assumed 2 days from onset to having a test + median 3.39 days for subsequent delays (Table 2)) we estimate that 26% (50% UI: 16, 40%; 95% UI: 7, 56%) of the secondary cases’ transmission potential occurs prior to tracing. Halving these delays by 50% leads to more than half of the pre-tracing transmission potential being averted, a decrease to 14% (50% UI: 9, 21%; 95% UI: 5, 42%). This halving therefore results in the 14-day quarantine with no test averting 82% (50% UI: 75, 87%; 95% UI: 53, 93%) of transmission potential overall (Figure 3).

**Figure 3:**
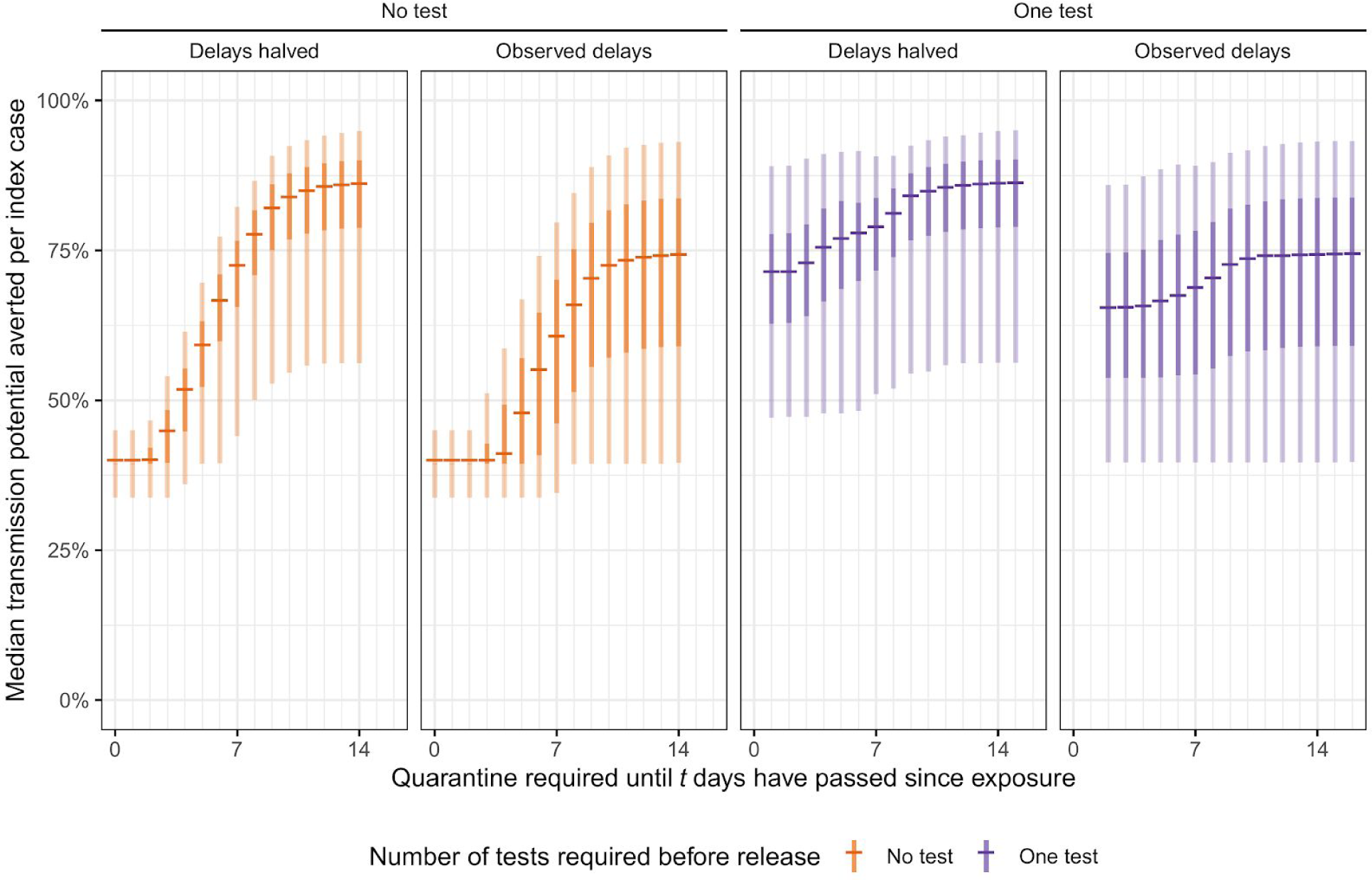
Transmission potential averted (integral of infectivity curve over time quarantined) in the no test, one test, and two test scenarios, with observed delays, and when delays are scaled down by 50%. Lines indicate the median amount of transmission potential averted for a given duration of quarantine, specified as the time since exposure to the index case. 95% and 50% uncertainty intervals are indicated by light and dark shaded areas, respectively

### Reduced adherence to quarantine

If adherence is imperfect, the ability of quarantine and testing to avert transmission potential is substantially reduced (Figure 4). For a 10% adherence rate to the 14-day post-exposure period, representingthe rate reported in the UK between March and August 2020 (4), 42% of transmission potential is averted (50% UI: 41, 43%; 95% UI: 35, 46%). A small, 10-percentage point increase in adherence raises the ability of shorter quarantines with testing to avert greater proportions of transmission potential compared to the current policy; however, adherence levels must be raised to above 40% to avert greater than 50% of median transmission potential for a 10 day quarantine with a test. If delays can be reduced by half, a greater proportion of transmission potential can be averted for a given level of adherence (Figure 4).

**Figure 4:**
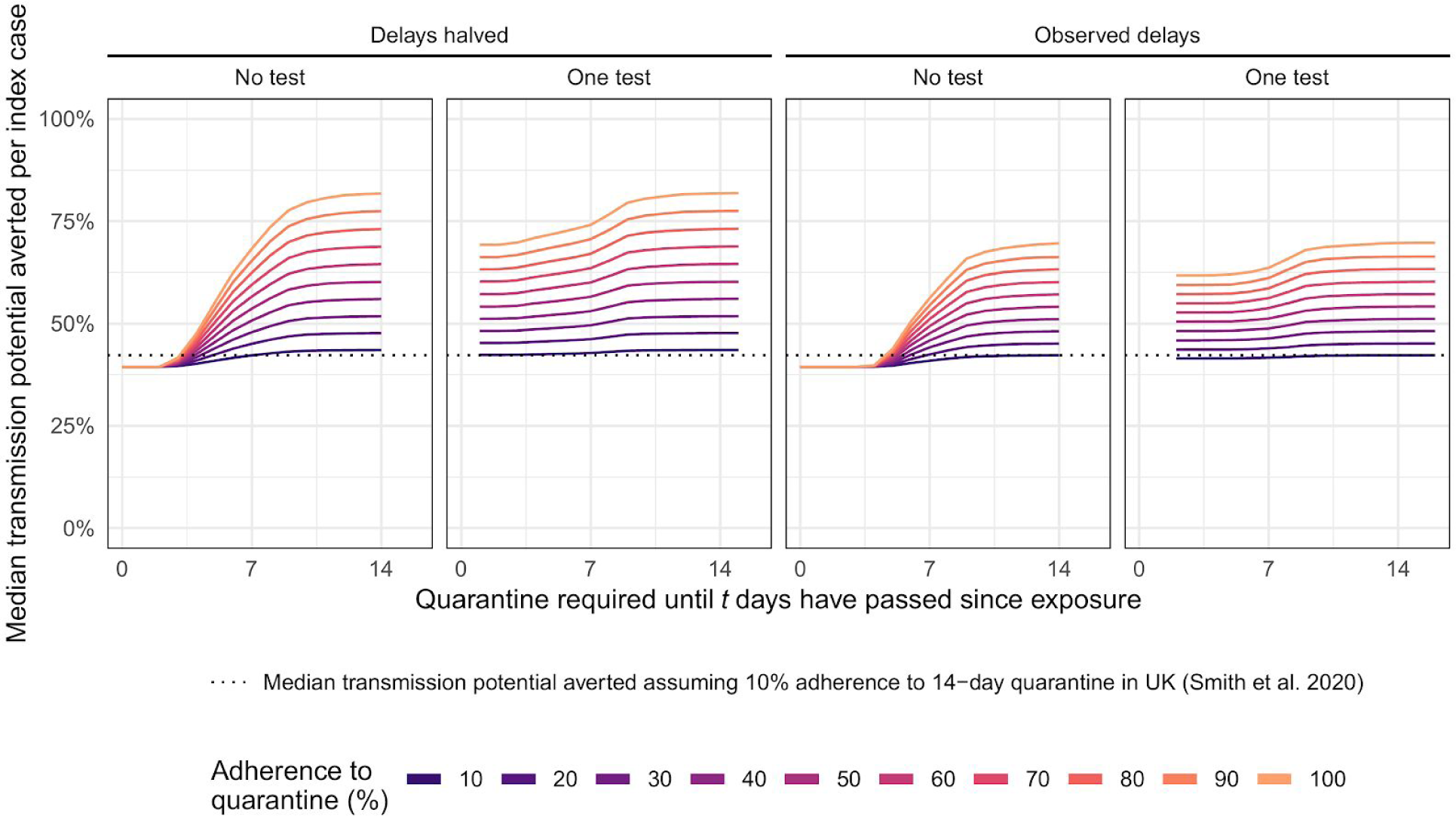
Median transmission potential averted, with adherence to quarantine varying from 10 to 100% adherence, in the no test, one test, and two test scenarios, with observed test and trace delays (median 3.9 days) and delays halved by 50%. Lines indicate the median amount of transmission potential averted for a given duration of quarantine, specified as the time since exposure to the index case. The dotted line indicates the median transmission potential averted assuming a 10.9% adherence rate to the 14-day quarantine period, as reported by Smith et al. 2020 (4).

## Discussion

Using a model combining SARS-CoV-2 natural history with reported contact tracing timings in the UK, we estimate the recommended 14 days of quarantine following last exposure from a confirmed case can prevent up to 70% (95% UI: 39, 90%) of onward transmission from secondary cases, assuming perfect adherence to quarantine. Similarly, a PCR test 7 days post-exposure with quarantine until a negative test result can prevent up to 68% (95% UI: 40, 88%) of tertiary cases. However, a test upon tracing may avert the majority (62%, 95% UI: 40, 84%) of transmission, which may represent a logistically viable strategy. The introduction of a PCR test at the end of quarantine mainly acts to detect and isolate still asymptomatic cases who would otherwise enter the community while still potentially infectious, assuming that symptomatic cases would self-isolate and hence not further transmit. However, the ability of a contact tracing programme to minimise the transmission potential of secondary cases is highly dependent on reducing the delays from the index cases’ onset to the tracing and quarantining of secondary cases and the adherence of secondary cases to quarantine guidelines for the duration of their quarantine. For example, we find that halving the delay from an index cases’ positive test result to tracing and quarantine of contacts to an average of 2 days (vs the current 4 days) translates to averting 82% (95% UI: 53, 93%) compared to 70% (95% UI: 39, 90%) in a 14-day quarantine, no-test scenario. Taking into account the reported low (∼10%) adherence rate to quarantine if contacted by test and trace in the UK (e.g, did not leave the house in the 14 day post-exposure period) (4), the median transmission potential averted at 14 days is 42% (95% UI: 35, 46%). As such, monitoring and reducing the delays in testing and tracing, and increasing adherence to quarantine, has a markedly greater impact on reducing onward transmission.

From late July to late August 2020 the average time to test results in satellite and home testing increased from approximately 36 to approximately 72 hours (8). Increasing delays due to pressure on testing throughput therefore represents a challenge to the effective operation of test and trace systems. From the fitted distribution of delays, approximately 25% of total times to complete tracing are greater than 5.1 days, the median incubation period for symptomatic infections. This runs the risk of a test and trace programme being minimally effective, as secondary cases may have been transmitting for a number of days in the community during the time the index case’s contact tracing is taking place. These delays mean that a “14 day quarantine” is in reality a 10 day quarantine (on average), due to the guidance that traced individuals isolate from the date of last exposure to the index case and not when they are traced. Factoring in the 2 day turnaround for test results in secondary cases means we assume that test-negative individuals taking a test 7 days post-exposure, for example, would leave quarantine on day 9. We have shown above that halving the delays in testing index cases and beginning contact tracing more than halves the transmission potential that occurs pre-tracing. In this analysis, we have focused on the potential for quarantine and testing to reduce the transmission potential of traced secondary infections and have not evaluated the number of tests which may be required, nor the possibility of false positives which despite the high specificity of PCR, may arise in mass testing of asymptomatic individuals.

In our analysis we assumed that delays in test result return, sourcing of contacts and contacting them to encourage quarantine are all independent. The delays may be positively correlated, however, with common structural causes, which could lead to a total delay distribution with a smaller median and larger variance. This would have the effect of getting more people into quarantine quicker, but those who are delayed in being quarantined are delayed longer. Longer delays to quarantining reduce the relative effectiveness of shorter quarantine periods in comparison to a 14 day quarantine period, as individuals spend less of their infectious period in quarantine after initial tracing and may exit prior to onset of symptoms. Longer delays also result in a predictable increase in the number of days individuals spend in the community with close to peak infectivity prior to being traced. Notably, the inclusion of PCR testing reduces the median transmission potential post-release to low levels (Figure S1). A no-test, 14-day quarantine period does not eliminate the risk of individuals spending time infectious after release. However, this represents individuals who are late in their infectious period and largely asymptomatic.

Requiring a PCR test provides some benefit with shorter quarantine periods. This additional benefit diminishes with longer quarantine duration, as infectious persons have a higher probability of developing symptoms (if ever-symptomatic) and self-isolating. The addition of a test on tracing has a negligible effect on reducing the transmission potential beyond that of a single test, and any additional benefit diminishes as the quarantine period increases in duration. An initial test upon tracing, if negative, may reduce the perceived importance of an individual needing to quarantine further, despite the high probability of the result being falsely negative at this early time point. However, testing traced secondary cases immediately may allow for rapid tracing of possible tertiary cases (i.e, infections generated from secondary cases), preventing additional chains of transmission. Additionally, while we have considered two independent PCR tests, future work should investigate the utility of regular (i.e weekly) testing with rapid, low-cost antigen or RT-LAMP tests. Due to the lower specificity of these methods, sequential testing with a follow-up PCR test conditional on a positive first test may also be considered.

We find uncertainty in our estimates of transmission potential primarily due to variation in symptom onset of secondary cases, the probability of detection by PCR, as well as variation in testing and tracing delays.

Due to a lack of currently available data, we have assumed that index cases effectively self-isolate (and hence cease generating secondary cases) once their symptoms develop to the point that they seek out and take a PCR test, with a central assumption of 2 days. However, if this period can be reduced through sensitisation of the public to COVID-19 symptoms and the importance of early action, shorter quarantine periods with testing at the end of the period becomes more viable. Digital contact tracing could improve the effectiveness of contact tracing through a reduction in the delays associated with sourcing and quarantining contacts(6), a process which we estimate currently takes an average of 3.9 days.

In this analysis we consider only the performance of quarantine and testing strategies with respect to infection history timings and tracing delays, and as such we do not consider other aspects of the test and trace system which may result in poor outcomes, such as the fraction of index cases that do not engage with the service (16), variation in the number of cases generated by each index case (17), or the proportion of secondary cases missed by tracers (18). A survey by Smith et al. found just 10.9% of individuals traced by NHS Track and Trace adhere to their quarantine completely (defined as did not leave the house for 14 days), indicating an inability or unwillingness to adhere to quarantine rules (19). Adherence to quarantine for other infectious diseases was found to be associated with greater trust in government; ease of understanding the specified quarantine protocol; perceived importance of quarantine in reducing transmission; strong social support networks; and the presence of income support or the provision of other supplies (20). It is possible that longer quarantine periods result in a decrease in both the proportion of individuals adhering to quarantine protocol and the degree to which they comply. Here we assumed perfect adherence as the baseline case with sensitivities based on constant but partial adherence between 10% and 90%, with comparison to the ∼10% rate reported by Smith et al. (4). However, applying this 10% uniformly to all individuals over their quarantine period may underestimate the transmission potential averted, as many reasons cited for breaking quarantine were activities of a short duration and low probability of contact with others (e.g, solo outdoor exercise or walking pets). We find that small increases in adherence may have a greater impact than introducing testing and reducing quarantine duration, although it is possible that introducing testing may alter the behaviour of quarantined individuals in both positive (individuals may adhere more strictly to quarantine if they are motivated by an upcoming tests) or negative (individuals may cease to self-isolate upon receipt of a negative initial test, if two tests are conducted) ways. Further work on COVID quarantine adherence is required in order to understand how quarantined individuals behave, and whether mandatory isolation of cases and suspected cases in hotels or hospitals may be necessary to prevent onward transmission.

We have shown that PCR testing combined with 7 days of quarantine could reduce the transmission potential from secondary cases notified through contact tracing to similar levels produced by a 14 day quarantine without testing, and that an initial test upon tracing may detect a majority of infections. However, factoring in structural issues in contact tracing such as testing and tracing delays and poor adherence of traced cases greatly reduces the ability of quarantine and testing to reduce onwards transmission, and addressing these should be a focus of policy. Future work should investigate the utility of rapid, frequent, and sequential testing, to aid in the detection and isolation of individuals infected with SARS-CoV-2.

## Data Availability

Model code is available at https://github.com/cmmid/pcr_track_trace

## Acknowledgements

This research was partly funded by the National Institute for Health Research (NIHR) (Billy Quilty is funded by 16/137/109 & 16/136/46) using UK aid from the UK Government to support global health research. The views expressed in this publication are those of the author(s) and not necessarily those of the NIHR or the UK Department of Health and Social Care. This research has been funded by UK Research and Innovation - MC_PC_19065 - Covid 19: Understanding the dynamics and drivers of the COVID-19 epidemic using real-time outbreak analytics (Samuel Clifford, W John Edmunds). This research was partly funded by the Wellcome Trust (Sir Henry Dale Fellowship: 208812/Z/17/Z, Stefan Flasche and Samuel Clifford; 206250/Z/17/Z, Adam J Kucharski). This project has received funding from the European Union’s Horizon 2020 research and innovation programme - project EpiPose (W John Edmunds).

The authors declare no conflicts of interest.

The following funding sources are acknowledged as providing funding for the working group authors. Alan Turing Institute (AE). BBSRC LIDP (BB/M009513/1: DS). This research was partly funded by the Bill & Melinda Gates Foundation (INV-001754: MQ; INV-003174: KP, MJ, YL; NTD Modelling Consortium OPP1184344: CABP, GFM; OPP1180644: SRP; OPP1183986: ESN; OPP1191821: KO’R, MA). BMGF (OPP1157270: KA). DFID/Wellcome Trust (Epidemic Preparedness Coronavirus research programme 221303/Z/20/Z: CABP, KvZ). DTRA (HDTRA1-18-1-0051: JWR). Elrha R2HC/UK DFID/Wellcome Trust/This research was partly funded by the National Institute for Health Research (NIHR) using UK aid from the UK Government to support global health research. The views expressed in this publication are those of the author(s) and not necessarily those of the NIHR or the UK Department of Health and Social Care (KvZ). ERC Starting Grant (#757699: JCE, MQ, RMGJH). This project has received funding from the European Union’s Horizon 2020 research and innovation programme - project EpiPose (101003688: KP, MJ, PK, RCB, YL). This research was partly funded by the Global Challenges Research Fund (GCRF) project ‘RECAP’ managed through RCUK and ESRC (ES/P010873/1: AG, CIJ, TJ). HDR UK (MR/S003975/1: RME). Nakajima Foundation (AE). NIHR (16/137/109: CD, FYS, MJ, YL; Health Protection Research Unit for Immunisation NIHR200929: NGD; Health Protection Research Unit for Modelling Methodology HPRU-2012-10096: TJ; NIHR200929: MJ; PR-OD-1017-20002: AR). Royal Society (Dorothy Hodgkin Fellowship: RL; RP\EA\180004: PK). UK DHSC/UK Aid/NIHR (ITCRZ 03010: HPG). UK MRC (LID DTP MR/N013638/1: GRGL, QJL; MC_PC_19065 - Covid 19: Understanding the dynamics and drivers of the COVID-19 epidemic using real-time outbreak analytics: AG, NGD, RME, TJ, YL; MR/P014658/1: GMK). Authors of this research receive funding from UK Public Health Rapid Support Team funded by the United Kingdom Department of Health and Social Care (TJ). Wellcome Trust (206250/Z/17/Z: TWR; 206471/Z/17/Z: OJB; 210758/Z/18/Z: JDM, JH, KS, NIB, SA, SFunk, SRM). No funding (AKD, AMF, CJVA, DCT, KEA, SH, YWDC).

## Supplementary appendix

### Reducing index cases’ test delays

In the case of a two day delay to seeking a test (single test with a two day turnaround at the end of the mandatory quarantine period), the amount of transmission potential averted with a 14 day quarantine is 70% (50% UI: 56, 80%; 95% UI: 40, 91%). By reducing the time to the index case’s test to one day, the transmission potential averted is increased to 74% (50% UI: 59, 84%; 95% UI: 40, 93%). An increase to 3 days’ delay results in 65% (50% UI: 53, 76%; 95% UI: 39, 87%) being averted.

**Figure S1:**
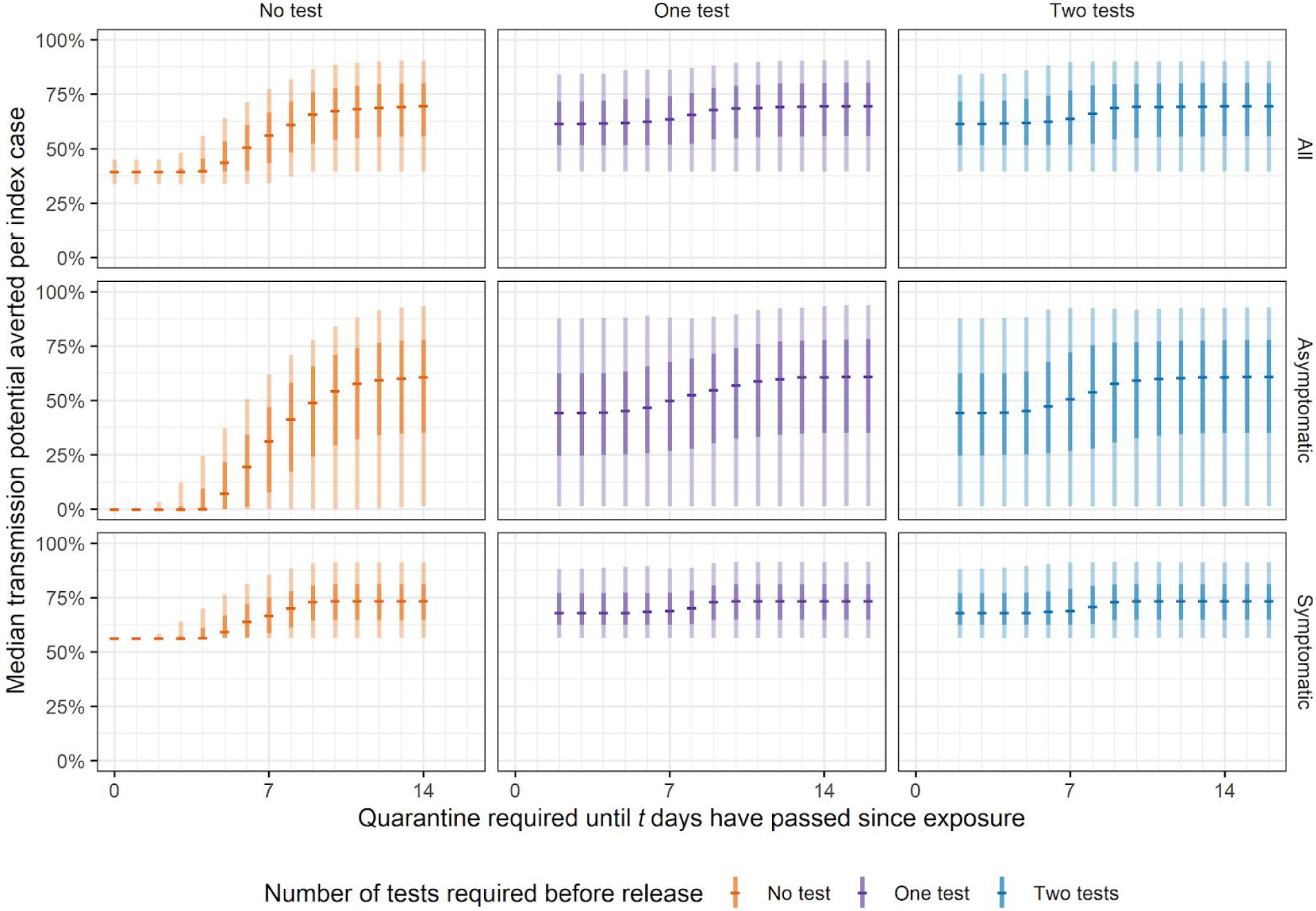
Transmission potential averted (integral of infectivity curve over time spent in quarantine and post-quarantine isolation) in the no test, one test, and two test scenarios, stratified by type of infection (all, asymptomatic, and symptomatic). Central bars indicate the median amount of transmission potential averted for a required time since exposure for which traced individuals must quarantine until. 95% and 50% uncertainty intervals indicated by light and dark shaded bars, respectively.

**Figure S2:**
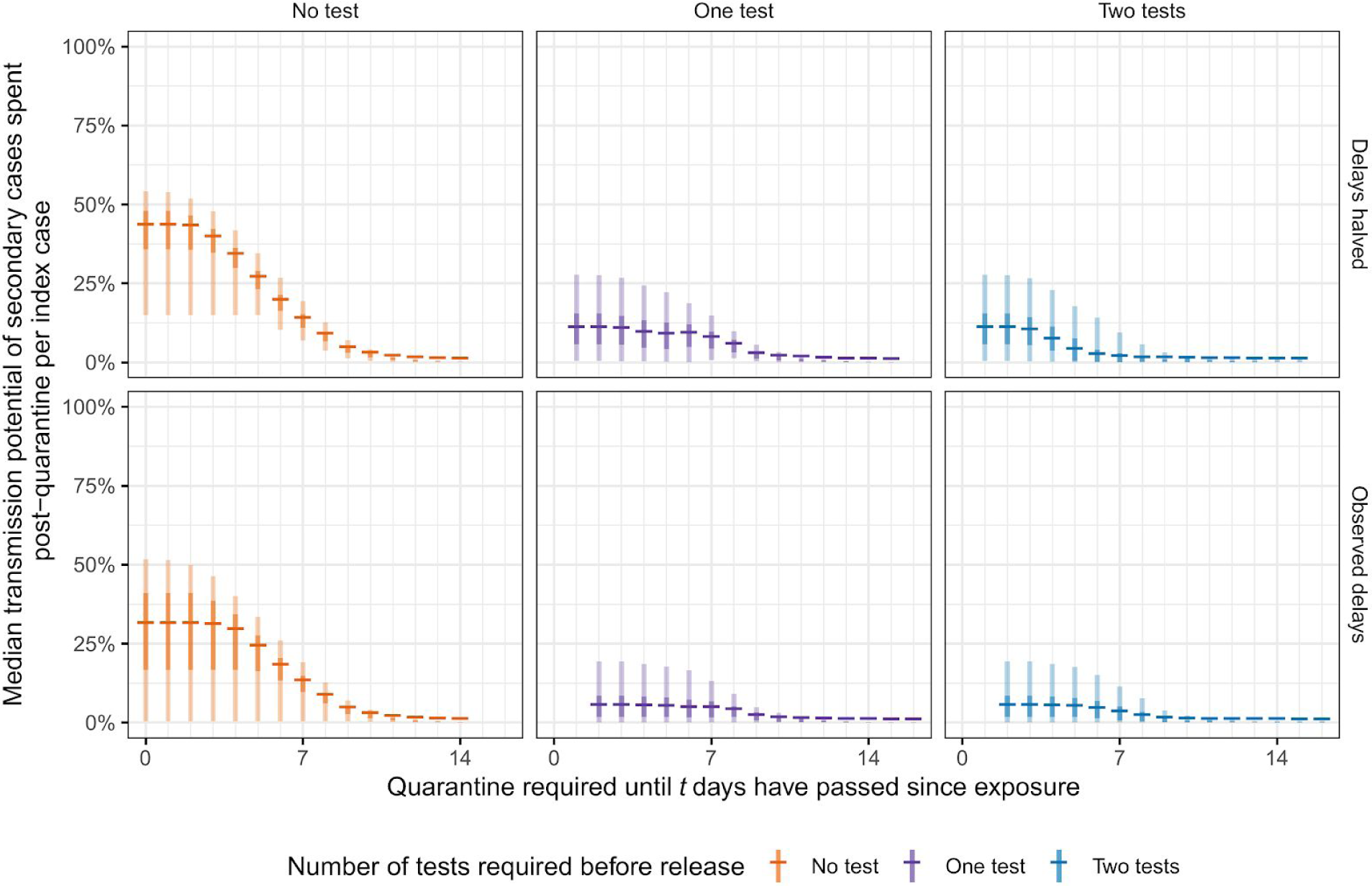
Transmission potential after leaving quarantine, stratified by number of tests. Lines indicate the median amount of transmission potential remaining after leaving quarantine for a given duration of quarantine, specified as the time since exposure to the index case. 95% and 50% uncertainty intervals indicated by light and dark shaded areas, respectively.

**Figure S3:**
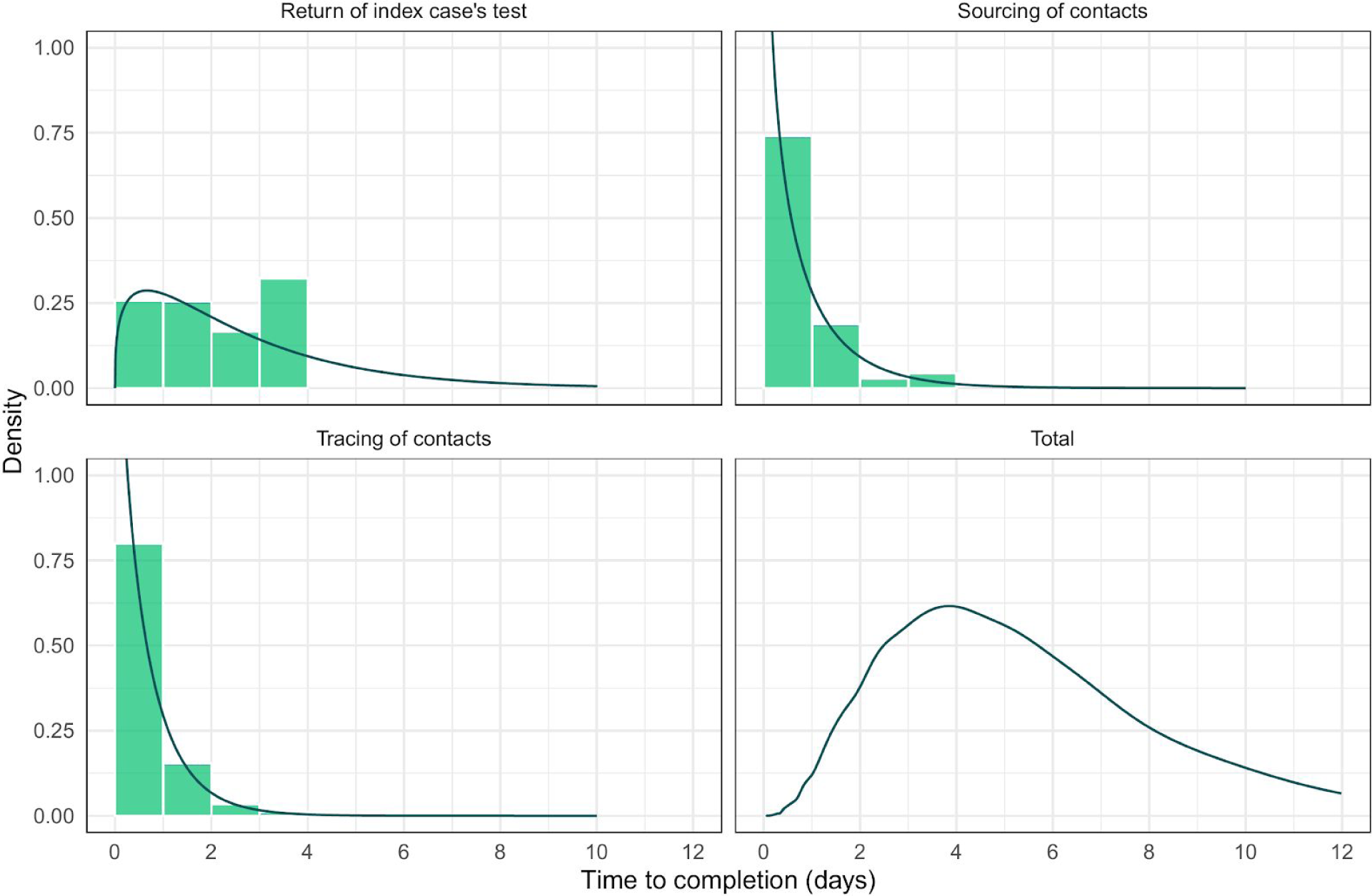
Observed delays (green bars) and modelled probability distributions (dark green curves) for the delays in the test and trace system. The total time delay is a kernel-smoothed empirical density based on the sum of 10,000 independent samples from the distributions fit to the observed delays.

**Figure S4:**
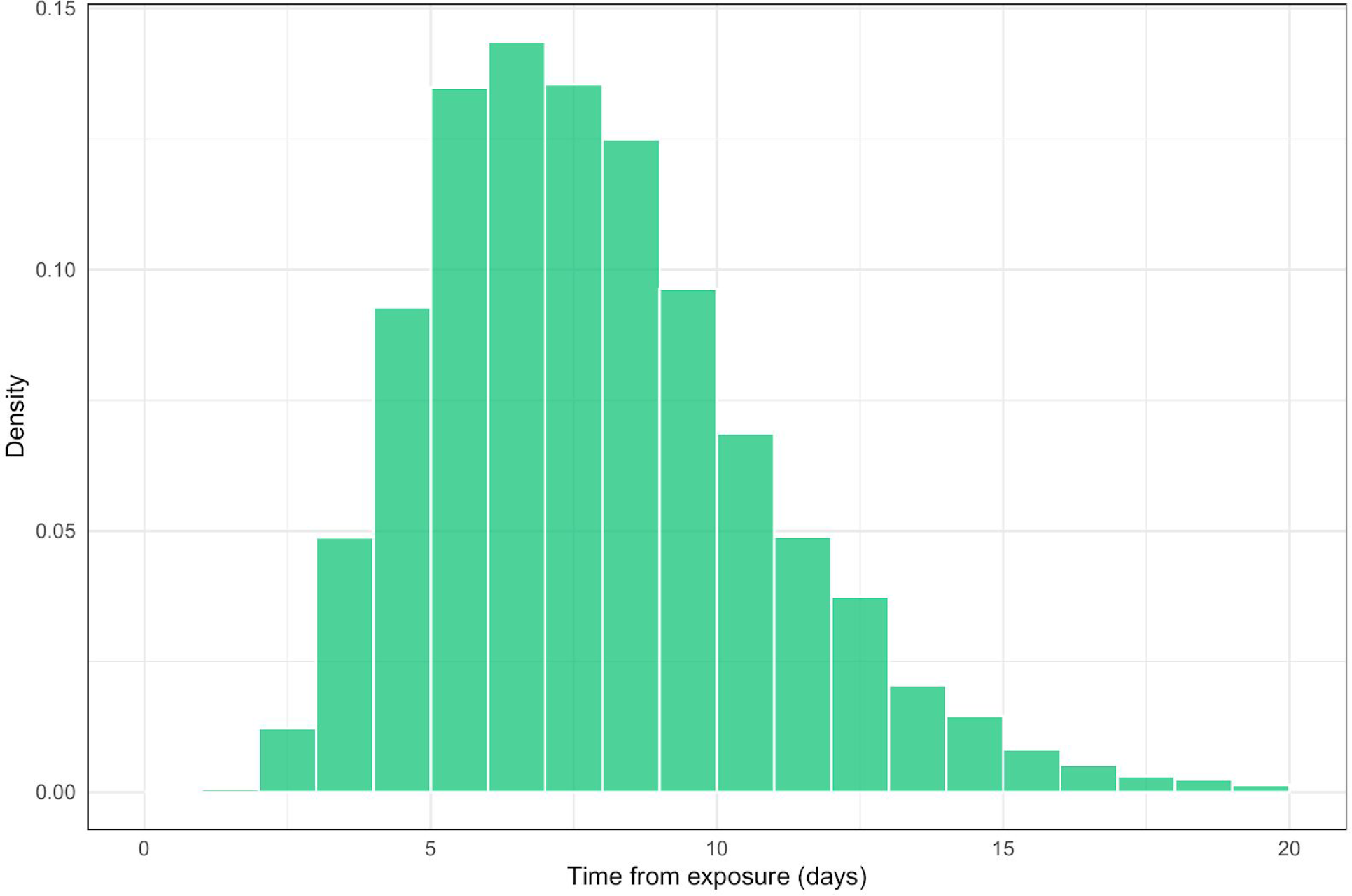
Transmission potential of persons infected with SARS-CoV-2, generated from the incubation period from Li et al. (2020) and infectivity profile from Ashcroft et al. (2020). Sampled sum of log-normally distributed onset of symptoms, with location parameter 1.63 and scale parameter 0.41, and Gamma-distributed infectivity from onset, with shape 97.19, rate 3.71, and shifted by 25.62 days.

**Figure S5:**
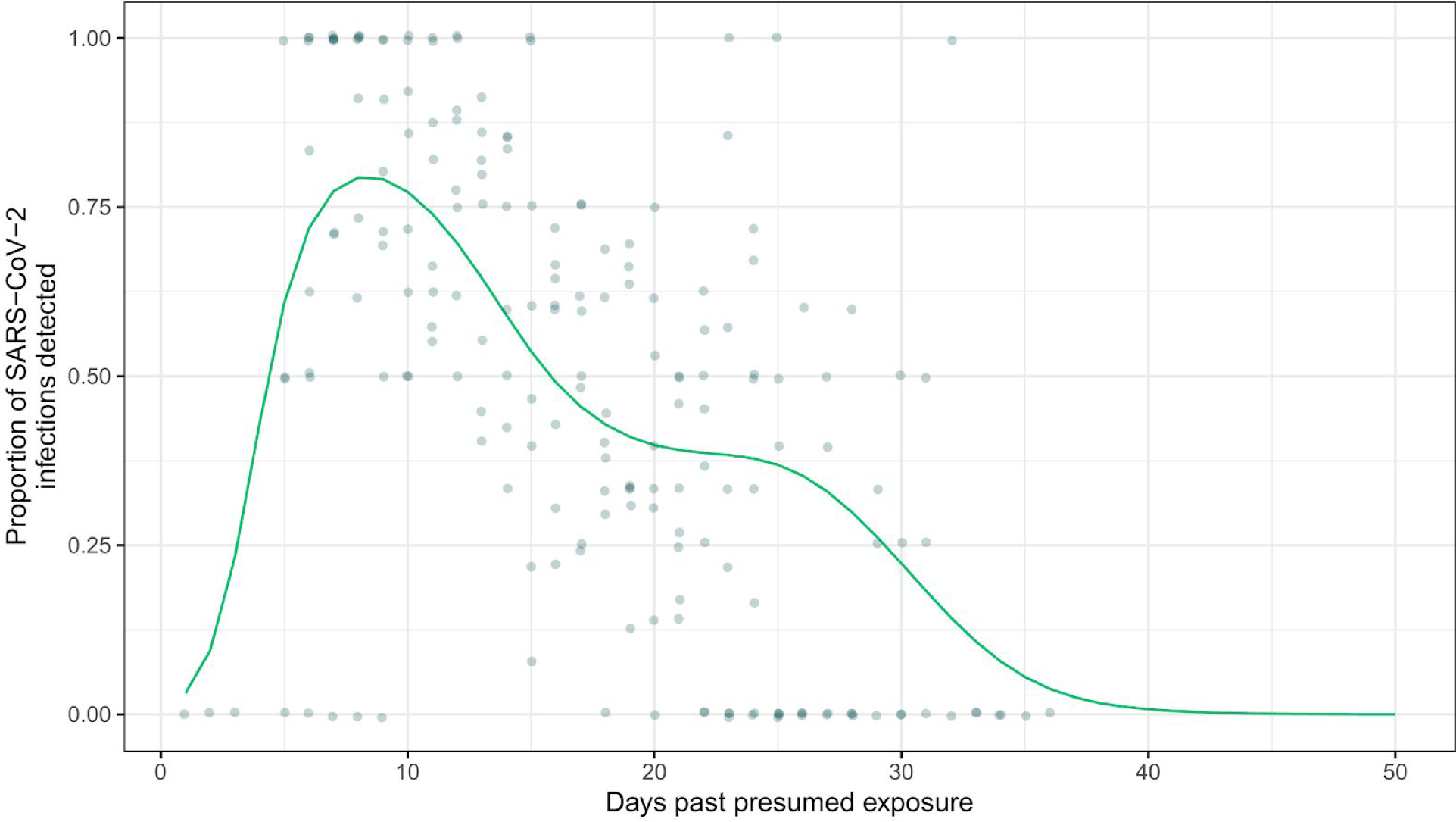
PCR sensitivity curves, obtained by fitting a Binomial GAM to the data collated in Kucirka et al. (2020) (13) The mean fit is used as the time-varying sensitivity function, *P* (*t*), and hence no uncertainty is shown in the figure.

**Figure S6:**
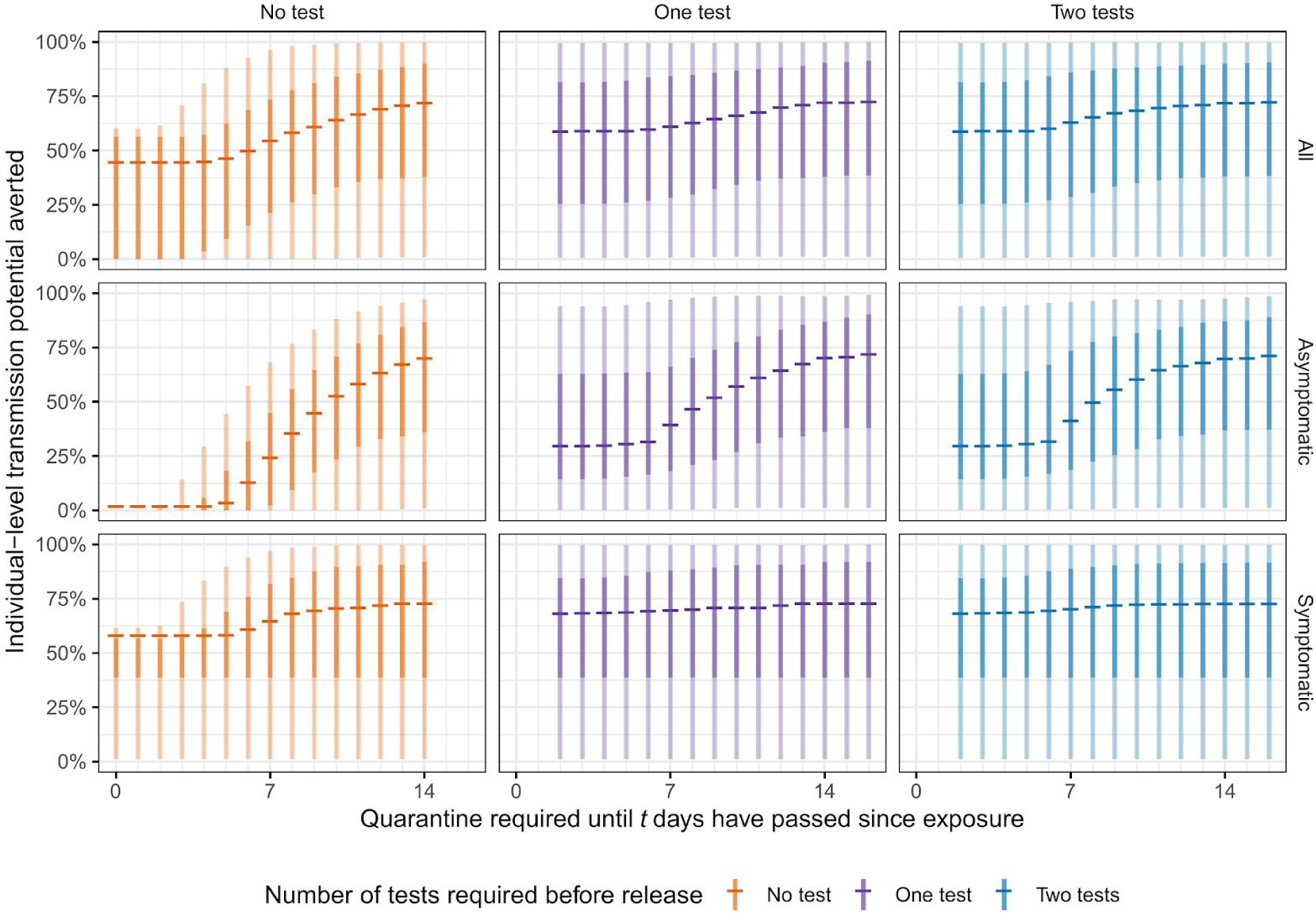
Per-secondary case transmission potential averted by quarantine and/or testing, stratified by number of tests, and symptomatic status. Generated by simulating 1000 index cases and 100 secondary cases per scenario. Central bars indicate the median amount of transmission potential remaining after leaving quarantine for a given duration of quarantine, specified as the time since exposure to the index case. 95% and 50% uncertainty intervals indicated by light and dark bars, respectively.

